# Postpartum hormonal contraceptive use and risk of depression

**DOI:** 10.1101/2024.09.27.24314424

**Authors:** Søren Vinther Larsen, Brice Ozenne, Anders Pretzmann Mikkelsen, Xiaoqin Liu, Kathrine Bang Madsen, Trine Munk-Olsen, Øjvind Lidegaard, Vibe G Frokjaer

**Author notes:** **Corresponding Author:** Søren Vinther Larsen, PhD, Neurobiology Research Unit, Copenhagen University Hospital - Rigshospitalet, building 8057, Blegdamsvej 9, DK-2100 Copenhagen, Denmark.

## Abstract

**Importance:** Hormonal contraceptive (HC) use is associated with depression. It is, however, unknown whether this is also true in the postpartum period where women have a heightened depression risk and are routinely offered to start HC.

**Objective:** To determine if HC initiation postpartum is associated with depression development within 12 months postpartum.

**Design:** A cohort study based on Danish register data.

**Setting:** Nationwide population-based.

**Participants:** All primiparous women in Denmark who gave birth from 1997 through 2022. Women were excluded if they had a depression within 24 months prior to delivery, had multiple birth or stillbirth, or a diagnosis of breast cancer or liver tumor.

**Exposure:** HC initiation within 12 months postpartum treated as a time-varying exposure. HC types were categorized as combined oral contraceptives (COCs), combined non-oral contraceptives (CNOCs), progestogen-only pills (POPs), and progestogen-only non-oral contraceptives (PNOCs).

**Main Outcome and Measure:** Depression defined as filling an antidepressant prescription or receiving a hospital depression diagnosis. Adjusted hazards ratios (HRs) and average absolute risks of depression within 12 months postpartum were estimated using Cox regression and a G-formula estimator.

**Results:** Of 610,038 first-time mothers, 41% initiated HC within 12 months postpartum (mean [SD] age; 27.6 [4.3] years for HC users vs. 29.6 [4.8] years for non-users). HC initiation was associated with subsequent depression with a HR of 1.49 (95% CI, 1.42;1.56) compared to no use resulting in an increase in the 12-month absolute risk from 1.36% (1.32;1.39) to 1.54% (1.50;1.57).The HR for COC was 1.72 (1.63;1.82); CNOC 1.97 (1.64;2.36); PNOC 1.40 (1.25;1.56). POP exposure was associated with an initially reduced instantaneous risk but it was increased late postpartum. Investigating the impact of timing of initiation showed that the earlier COC was initiated postpartum the higher the associated rate ratio of depression.

**Conclusions and Relevance:** HC initiation postpartum was associated with 49% higher instantaneous depression risk resulting in an increase in 12-month risk from 1.36% to 1.54% in the observed population. Such associated risk was higher the earlier it was initiated postpartum, at least for COC. This raises the question if depression incidence postpartum is inflated by routine HC initiation after childbirth.

## Introduction

The postpartum period is a critical time to avoid unintended and unwanted pregnancies, as short interpregnancy intervals may lead to increased perinatal and maternal health risks.^1,2^ In Denmark, as many as 40% of mothers initiate hormonal contraception (HC) within the first year after delivery and throughout the last 20 years they have started sooner and sooner after delivery.^3^

HC initiation has been associated with an increased risk of developing depression, especially in adolescents.^4–6^ It is unclear whether this also applies in the postpartum period,^7^ where women are already at heightened risk of developing depression.^8^ This has been hypothesized to be, at least in some women, attributed to the large hormonal withdrawal after the delivery.^9^ This drastic hormonal shift is incomparable to any other lifetime events, hence women who recently gave birth may be differently affected by exogenous hormone exposure, than women at other lifetime periods. This raises the question whether the routine practice of HC initiation in the postpartum period inflates the already heightened risk of depression. Only a few studies have previously addressed this and found conflicting results, however, they were limited by lack of generalizability, insufficient follow-up time, and insufficiently accounting for potential confounders.^7^

Here, we take advantage of the Danish national health registers to investigate the link between HC initiation postpartum and depression risk in a large, unselected population with one year follow-up time while accounting for various potential confounders such as medical indications for HC use. Specifically, the objective is to determine if HC initiation postpartum increases the risk of depression in the postpartum period compared to no HC exposure and whether it depends on age, HC type and timing of initiation postpartum.

## Methods

### Study design

This population-based cohort study used healthcare data from the registers listed in **eTable 1**. Data were linked via the unique personal identification number given to Danish residents at birth or immigration. The data were provided by the Danish e-Health Authority and approval was obtained from the Regional Data Health Board “Privacy”. No ethical approval or informed consent are required for register-based studies in Denmark. The study was reported according to the Strengthening the Reporting of Observational Studies in Epidemiology guidelines.^10^

### Study population

The source population included all primiparous women living in Denmark for at least 24 months before giving birth between January 1^st^ 1997 through December 2022 (n=663,654), identified from the Danish Civil Registration System and the Medical Birth Registry.^11,12^ We excluded 34,633 women who had received a depression diagnosis or filled a prescription for antidepressant medication within 24 months before delivery to ensure only new cases of depression onset were included, 16,316 women who had a multiple birth or stillbirth,^13,14^ and 2,667 women who had a contraindication for any HC type in form of prior diagnosis of breast cancer or a liver tumor.^15^ A total of 610,038 first-time mothers were left in the analyses (**eFigure 1**).

### Exposure

HC use postpartum was treated as a time-varying exposure such that all women contributed to non-exposed time until the day they filled a HC prescription after giving birth and hereafter contributed to the exposed time for the rest of the follow-up time. The exposure was differentiated by the following HC types; combined oral contraceptives (COCs); combined non-oral contraceptives (CNOCs) (patch and vaginal ring); progestogen-only pills (POPs); and progestogen-only non-oral contraceptives (PNOCs) (implant, depot injection, and levonorgestrel-releasing intrauterine system (LNG-IUS)).

### Outcome

The outcome of interest was depression within 12 months after delivery. Depression was defined as filling a prescription of antidepressant medication identified in the National Prescription Register (ATC: N06A*),^16^ or a hospital discharge diagnosis of depression identified in the National Patient Register (ICD-10: F32-34, F38, F39, F530).^17^

### Covariates

The following covariates measured at time of delivery were included in the analyses: Maternal age (below 20 years, 20-29 years, and 30 years and above); highest educational level (below high school, high school or vocational education, or bachelor degree or above); civil status (married/registered with a partner or not); history of any other mental disorder including previous use of psychoactive drugs; parental history of mental disorders; medical indications for HC use including polycystic ovary syndrome, endometriosis, premenstrual syndrome, dysmenorrhea, heavy menstrual bleeding, hirsutism, and acne; having received in vitro fertilization treatment; preterm birth; instrument-assisted- or cesarean delivery; pre-eclampsia/eclampsia; pre-gestational- or gestational diabetes; and finally, we controlled for period effects by including calendar-year in 5-year bands.

### Statistical analysis

The mothers were followed from day of delivery until 12 months postpartum, the development of depression, emigration, death, or end of the study period, whichever came first. Death being very rare (<0.01%), we only modelled the hazard of depression and neglected the hazard of death when computing risks and average risks. We used Cox proportional hazard models adjusted for the listed covariates to estimate hazard ratios (HRs) for developing depression in the postpartum period between HC exposed vs. non-exposed. To examine whether the association differed by HC type and age group, we further analyzed the association in two Cox models: one stratified by HC type and another stratified by age group. In addition, we applied a G-formula to the estimates from a multistate Cox model^18^ to quantify the average estimated risk of developing depression in the postpartum period under the observed HC use as well as in the hypothetical situation where nobody started HC (see **eMethods 1** and **eFigure 2** for details). The risk difference between these two scenarios is referred to as the effect of HC on depression in the postpartum period in the investigated population.

Exploratively, we investigated the effect of postpartum timing of COC initiation on the risk of depression (we were not able to reliably test this for the other types due to limited sample sizes). As both time since delivery^19^ and time since COC initiation^4^ may influence the depression risk, we included both time scales in a flexible parametric multistate model (see **eMethods 2** for details).^20^

To assess the proportionality hazards assumption for the exposure, we considered a Cox model with time-varying coefficient for the exposure using the *cox.aalen* function of the *timereg* package.^21^ The exposure showing violation of the proportion hazards assumption was modelled with a flexible parametric survival model via the *stpm2* function of the *rstpm2* package to display the hazard ratio as a function of time.^22^ To assess the sensitivity of the estimates to the proportional hazards assumption on the covariates, we used a test based on Schoenfeld residuals to identify possible violation of the proportional hazards assumption. The baseline hazard of the Cox regression model was stratified on the variables showing evidence for non-proportionality.

### Sensitivity analyses

We conducted five sensitivity analyses; first, we adjusted for immigration status (immigrant or descendant of an immigrant), smoking status, and highest obtained educational level of the parents. Due to considerable missing information about the parents’ educational level (7.8%) and smoking status (7.9%), we imputed missing values based on the listed covariates, exposure, and outcome with the *MICE* package ^23^ where binary variables were imputed with logistic regression and categorical data with polytomous logistic regression with 10 iterations and five imputations. Second, we re-defined the PNOC group to only include LNG-IUS to reduce the risk of confounding by indication. Third, a 28-day delay in HC exposure was used to account for possible delays in HC initation. Fourth, we right-censored women if they became pregnant within the follow-up time. Last, we stratified the association between HC exposure and depression on prior mental disorders, as studies suggest it moderates the association between HC use and depression risk.^24,25^

All analyses were conducted using R version 4.2.2.^26^ The analysis plan was pre-registered on www.aspredicted.gov (#125589).

## Results

Of 610,038 first-time mothers included in the study, 248,274 (40.7%) started using HC, of which 143,751 (23.6%) initiated COC, 5,465 (0.9%) CNOC, 66,612 (10.9%) POP, and 32,446 (5.3%) PNOC within the first year after delivery. 48 (0.008%) women died within 12 months after delivery and 3,287 (0.54%) were lost to follow-up due to emigration. Their characteristics and clinical profiles are shown in **Table 1**. The timing of initiation of the different types is illustrated by cumulative incidence curves in **eFigure 3A-B**. Notably, about 50% of those who initiated POP filled their first prescription between seven to 10 weeks postpartum.

**Table 1.** Characteristics and clinical profiles of users of different types of hormonal contraception and of non-users.

|  |  |  |  |  |  |  |  |
| --- | --- | --- | --- | --- | --- | --- | --- |
| **Profile** | **Exposure, No. (%)** |  |  |  |  |  |  |
|  | **Non-users** | **HC users** | **COC users** | **CNOC users** | **POP users** | **PNOC users** |  |
|  | **No. (%)** | **No. (%)** | **No. (%)** | **No. (%)** | **No. (%)** | **All users**<br>**No. (%)** | **IUS users**<br>**No. (%)** |
| Total | 361,764 (59.3) | 248,274 (40.7) | 143,751 (23.6) | 5465 (0.9) | 66,612 (10.9) | 32,446 (5.3) | 29,864 (4.9) |
| **Maternal age, y** |  |  |  |  |  |  |  |
| <20 | 6429 (1.8) | 8426 (3.4) | 6108 (4.2) | 323 (5.9) | 1039 (1.6) | 956 (2.9) | 480 (1.6) |
| 20-29 | 194,036 (53.6) | 171,993 (69.3) | 104,171 (72.5) | 3720 (68.1) | 44,203 (66.4) | 19,899 (61.3) | 18,122 (60.7) |
| ≥30 | 161,299 (44.6) | 67,855 (27.3) | 33,472 (23.3) | 1422 (26.0) | 21,370 (32.1) | 11,591 (35.7) | 11,262 (37.7) |
| **Educational level<sup>a</sup>** |  |  |  |  |  |  |  |
| Below high school | 47,340 (13.1) | 42,448 (17.1) | 29,817 (20.7) | 1227 (22.5) | 7395 (11.1) | 4009 (12.4) | 2554 (8.6) |
| High school/vocational education | 123,022 (34.0) | 103,969 (41.9) | 67,357 (46.9) | 2120 (38.8) | 24,920 (37.4) | 9572 (29.5) | 8816 (29.5) |
| Bachelor degree or above | 187,427 (51.8) | 100,777 (40.6) | 45,951 (32.0) | 2075 (38.0) | 34,007 (51.1) | 18,744 (57.8) | 18,412 (61.7) |
| **Highest educational level of parents<sup>b</sup>** |  |  |  |  |  |  |  |
| Below high school | 50,562 (14.0) | 40,258 (16.2) | 28,407 (19.8) | 759 (13.9) | 8059 (12.1) | 3033 (9.3) | 2361 (7.9) |
| High school/vocational education | 145,299 (40.2) | 120,518 (48.5) | 73,409 (51.1) | 2583 (47.3) | 30,619 (46.0) | 13,907 (42.9) | 12,622 (42.3) |
| Bachelor degree or above | 129,051 (35.7) | 77,059 (31.0) | 37,021 (25.8) | 1814 (33.2) | 24,340 (36.5) | 13,884 (42.8) | 13,422 (44.9) |
| Familial disposition for mental disorder | 36,859 (10.2) | 27,869 (11.2) | 14,254 (9.9) | 759 (13.9) | 8214 (12.3) | 4642 (14.3) | 4040 (13.5) |
| History of mental disorder | 65,720 (18.2) | 44,382 (17.9) | 22,213 (15.5) | 1275 (23.3) | 12,980 (19.5) | 7914 (24.4) | 7015 (23.5) |
| Immigrant or descendant of immigrant | 55,378 (15.3) | 18,606 (7.5) | 9055 (6.3) | 514 (9.4) | 6334 (9.5) | 2703 (8.3) | 2334 (7.8) |
| Civil status | 140,647 (38.9) | 70,553 (28.4) | 40,693 (28.3) | 1433 (26.2) | 19,549 (29.3) | 8878 (27.4) | 8467 (28.4) |
| Smoking status<sup>c</sup> | 40,006 (11.1) | 39,414 (15.9) | 27,479 (19.1) | 1024 (18.7) | 7342 (11.0) | 3569 (11.0) | 2620 (8.8) |
| Medical indication for HC | 19,062 (5.3) | 10,161 (4.1) | 5145 (3.6) | 259 (4.7) | 2944 (4.4) | 1813 (5.6) | 1689 (5.7) |
| IVF treatment | 64,073 (17.7) | 14,260 (5.7) | 7587 (5.3) | 292 (5.3) | 4234 (6.4) | 2147 (6.6) | 2074 (6.9) |
| (Pre-)gestational diabetes | 11,634 (3.2) | 6677 (2.7) | 3447 (2.4) | 133 (2.4) | 1895 (2.8) | 1202 (3.7) | 1079 (3.6) |
| Eclampsia/preeclampsia | 15,895 (4.4) | 11,376 (4.6) | 6484 (4.5) | 225 (4.1) | 3058 (4.6) | 1609 (5.0) | 1467 (4.9) |
| Preterm birth<sup>d</sup> | 21,206 (5.9) | 15,608 (6.3) | 9513 (6.6) | 308 (5.6) | 3984 (6.0) | 1803 (5.6) | 1605 (5.4) |
| Instrument-assisted delivery | 121,217 (33.5) | 83,345 (33.6) | 49,471 (34.4) | 1860 (34.0) | 22,173 (33.3) | 9841 (30.3) | 9019 (30.2) |

<sup>a</sup>0.4%, 0.4%, 0.8%, 0.4%, 0.4%, 0.3%, and 1.1% had missing information among mothers initiating HC, COC, CNOC, POP, PNOC, IUS, and not initiating HC postpartum, respectively.
<sup>b</sup>4.2%, 3.4%, 5.7%, 5.4%, 5.0%, 4.9%, and 10.2% had missing information among mothers initiating HC, COC, CNOC, POP, PNOC, IUS, and not initiating HC postpartum, respectively.
<sup>c</sup>6.9%, 9.4%, 2.5%, 4.0%, 2.8%, 2.8%, and 8.6% had missing information among mothers initiating HC, COC, CNOC, POP, PNOC, IUS, and not initiating HC postpartum, respectively.
<sup>d</sup>0.7%, 1.0%, 0.5%, 0.4%, 0.3%, 0.3%, and 1.3% had missing information among mothers initiating HC, COC, CNOC, POP, PNOC, IUS, and not initiating HC postpartum, respectively.
HC, hormonal contraceptive. aHR, adjusted hazard ratio. COC, combined oral contraceptive. CNOC, combined non-oral contraceptive. POP, progesterone-only pill. PNOC, progesterone-only non-oral contraceptive. IUS, intrauterine system. IVF, in vitro fertilization.

Within 12 months after delivery, 9251 (1.5%) of the first-time mothers developed a depression. The crude incidence rate of depression was 21 per 1,000 person-years for the mothers exposed to HC and 14 per 1,000 person-years for non-exposed mothers resulting in an adjusted HR of 1.49 (95% CI, 1.42;1.56) (**Table 2**).

**Table 2.** Hazard ratios for depression in the postpartum period for HC exposed compared to non-exposed mothers.

|  |  |  |  |  |  |
| --- | --- | --- | --- | --- | --- |
| Exposure | Person-years | No. of events | aHR<sup>a</sup> (95% CI) | aHR<sup>b</sup> (95% CI) | aHR<sup>c</sup> (95% CI) |
| Non-exposed | 434 620 | 5980 | 1 (reference) | 1 (reference) | 1 (reference) |
| HC exposed | 156 928 | 3271 | 1.53 (1.46;1.61) | 1.49 (1.42;1.56) | 1.46 (1.40;1.53) |
| COC | 83 246 | 2157 | 1.83 (1.73;1.93) | 1.72 (1.63;1.82) | 1.68 (1.59;1.77) |
| CNOC | 3334 | 121 | 2.20 (1.84;2.64) | 1.97 (1.64;2.36) | 1.93 (1.61;2.31) |
| POP<sup>d</sup> | 48 886 | 640 | See eFigure 4 |  |  |
| PNOC | 21 462 | 353 | 1.42 (1.27;1.58) | 1.40 (1.25;1.56) | 1.38 (1.23;1.54) |
| IUS | 19 722 | 272 | 1.22 (1.08;1.38) | 1.27 (1.12;1.44) | 1.26 (1.11;1.43) |

<sup>a</sup>Adjusted for age and calendar year.
<sup>b</sup>Adjusted for age, calendar year, educational level, civil status, history of mental disorder, parental disposition for mental disorders, medical indications for HC use, IVF-treatment, preterm birth, instrument-assisted- or caesarean delivery, pre-eclampsia/eclampsia, pre-gestational- or gestational diabetes. Missing values were handled as a separate group for educational level and preterm delivery status.
<sup>c</sup>Adjusted for age, calendar year, educational level, civil status, history of mental disorder, parental disposition for mental disorders, medical indications for HC use, IVF-treatment, preterm birth, instrument-assisted- or caesarean delivery, pre-eclampsia/eclampsia, pre-gestational- or gestational diabetes, immigration status, parental educational level, and smoking status. Missing values were handled by use of multiple imputations for educational level, parental education, smoking status, and preterm delivery status.
<sup>d</sup>HRs for POP violated the non-proportional hazard assumption, hence the time-varying HR is shown in **Figure 1**.
HC, hormonal contraceptive. aHR, adjusted hazard ratio. COC, combined oral contraceptive. CNOC, combined non-oral contraceptive. POP, progesterone-only pill. PNOC, progesterone-only non-oral contraceptive. IUS, intrauterine system.

When stratified on HC type, the adjusted HR was 1.72 (1.63;1.82) for COC, 1.97 (1.64;2.36) for CNOC, 1.40 (1.25;1.56) for PNOC, and specifically for LNG-IUS it was 1.27 (1.12;1.44). As POP exposure showed time-varying effects, the HR for POP is represented as a function of time in **eFigure 4**, which shows a HR<1 early postpartum, which subsequently increased across the postpartum period to be significantly increased from about eight months postpartum. For mothers younger than 20 years of age, HC initiation postpartum was associated with depression with an HR of 1.62 (1.37;1.93), which was 1.55 (1.46;1.64) for mothers aged 20-29 years and 1.35 (1.24;1.47) for mothers aged 30 years or older (**eTable 2**). Adjustment for immigration- and smoking status and highest obtained educational level of the parents led to very similar estimated HRs (maximum absolute difference of 0.04) (**Table 2, eTable2**).

When the proportional hazards assumption was relaxed for covariates, for which there was evidence indicating violation of the proportional hazard, the estimates did not change remarkably (largest absolute difference was 0.11 for CNOC), except the estimated HR for the mothers younger than 20 years that were considerably lower (1.34 instead of 1.62) (**eFigure 5**).

Sensitivity analyses showed similar results (**eFigure 6**), but notably the HR was 1.63 (1.53;1.73) for women with no prior mental disorder (n=499,936) and 1.32 (1.23;1.41) for women with prior mental disorder (n=110,102), and the ratio between the two was 1.24 (1.13;1.35).

The average absolute risk of depression at 12 months postpartum for the study population under the observed HC use was 1.54% (1.50;1.57) (**Table 3**). In comparison, in the hypothetical scenario had no one initiated HC, the estimated average risk was 1.36% (1.32;1.39) (**Figure 1A**), resulting in a risk difference of 0.18% (0.16;0.20) (**Figure 1B**). Had all those who initiated a HC started on COC, the estimated average risk would have been 1.62% (1.58;1.65), on CNOC 1.70% (1.58;1.83), on POP 1.37% (1.33;1.41), and on PNOC 1.50% (1.44;1.56) with a risk of 1.45% (1.40;1.52) specifically for LNG-IUS (**Table 3**, **Figure 1C-D**). When the proportional hazard assumption was relaxed, the estimated risks were similar, but the confidence intervals were typically wider (**eTable 3** and **eFigure 7**).

**Table 3.** 12-month average absolute risk of depression in the postpartum period.

|  |  |  |
| --- | --- | --- |
| **Scenario** | **Absolute risk<sup>a</sup>**<br>**% (95% CI)** | **Absolute risk difference<sup>a</sup>**<br>**% (95% CI)** |
| No HC initiation<sup>b</sup> | 1.36 (1.32;1.39) | Reference |
| Observed HC initiation | 1.54 (1.50;1.57) | 0.18 (0.16;0.20) |
| All initiating HC start on COC<sup>c</sup> | 1.62 (1.58;1.65) | 0.26 (0.23;0.29) |
| All initiating HC start on CNOC<sup>c</sup> | 1.70 (1.58;1.83) | 0.35 (0.23;0.47) |
| All initiating HC start on POP<sup>c</sup> | 1.37 (1.33;1.41) | 0.01 (-0.02;0.04) |
| All initiating HC start on PNOC<sup>c</sup> | 1.50 (1.44;1.56) | 0.14 (0.09;0.20) |
| All initiating HC start on IUS<sup>c</sup> | 1.45 (1.40;1.52) | 0.10 (0.04;0.16) |

<sup>a</sup>Adjusted for age, calendar year, educational, civil status, immigration status, history of mental disorder; parental disposition for mental disorders, medical indications for HC use, IVF-treatment, preterm birth, instrument-assisted- or caesarean delivery, pre-eclampsia/eclampsia, pre-gestational- or gestational diabetes. Missing values were handled by imputing to separate groups for educational level and preterm delivery status.
<sup>b</sup>A hypothetical scenario obtained by setting the transition hazard from non-exposed to HC exposed to 0.
<sup>c</sup>A hypothetical scenario obtained by setting the transition hazard from non-exposed to COC/ CNOC/ POP/ PNOC/ IUS by adding the transition hazard for each type and by setting the transition hazard to 0 for the rest of the types. HC, hormonal contraceptive. COC, combined oral contraceptive. CNOC, combined non-oral contraceptive. POP, progesterone-only pill. PNOC, progesterone-only non-oral contraceptive. IUS, intrauterine system.

**Figure 1.**
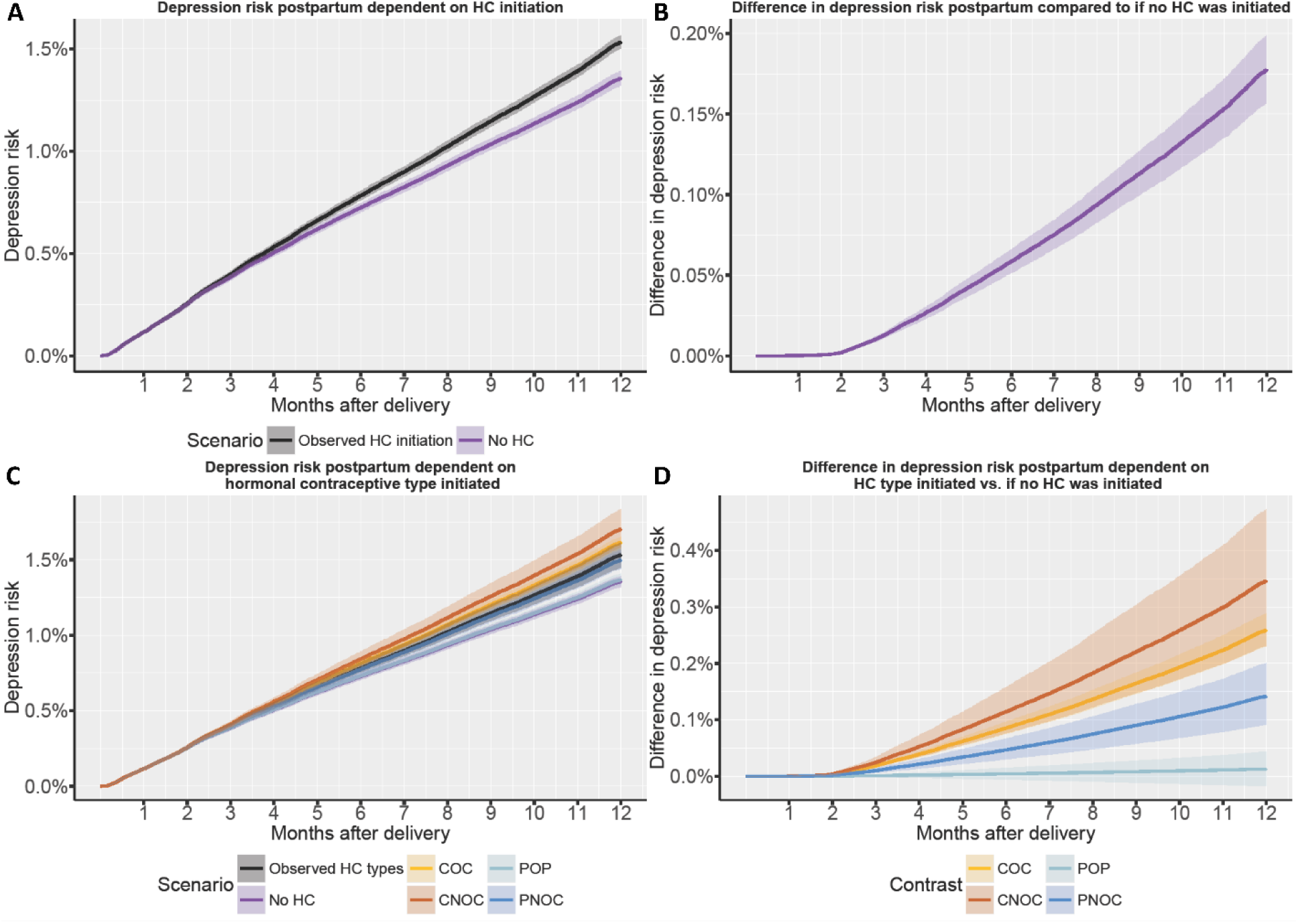
**A)** Average absolute risk with 95% confidence intervals of depression within 12 months from delivery under scenario 0; observed initiation of hormonal contraception (HC) postpartum vs. scenario 1; nobody had initiated HC. **B)** Average absolute risk difference with 95% confidence interval within 12 postpartum between these two scenarios. **C**) Average absolute risk with 95% confidence intervals of depression within 12 months from delivery for the scenarios where all mothers who are observed to initiate HC initiated either only combined oral contraception (COC, scenario 2), combined non-oral contraception (CNOC), progestogen-only pill (POP), or progestogen-only non-oral contraception (PNOC), with the observed transition intensity observed for each type vs. if nobody had initiated HC. **D**) Average absolute risk difference with 95% confidence interval within 12 postpartum contrasting scenarios 0 to scenario 2-5.

Exploratively, we found no evidence of an effect of time since initiation of COC on depression rate (**eFigure 8**), hence the effect of timing of initiation was modelled without including time since initiation as time scale. Early postpartum COC initiaton was associated with a higher rate of depression which gradually decreased during the first 7 months compared to no use (**Figure 2**, **eFigure 9**). The rate ratio remained above one during the whole time period, however, after 7 months it was estimated with high uncertainty due to the short remaining follow-up time. Likelihood-ratio tests showed evidence of a negative linear effect of time to COC initiation on depression rate, i.e., the curve in **Figure 2** can be summarized by a rate ratio (per year after delivery) of 0.61 (0.48;0.79).

**Figure 2.**
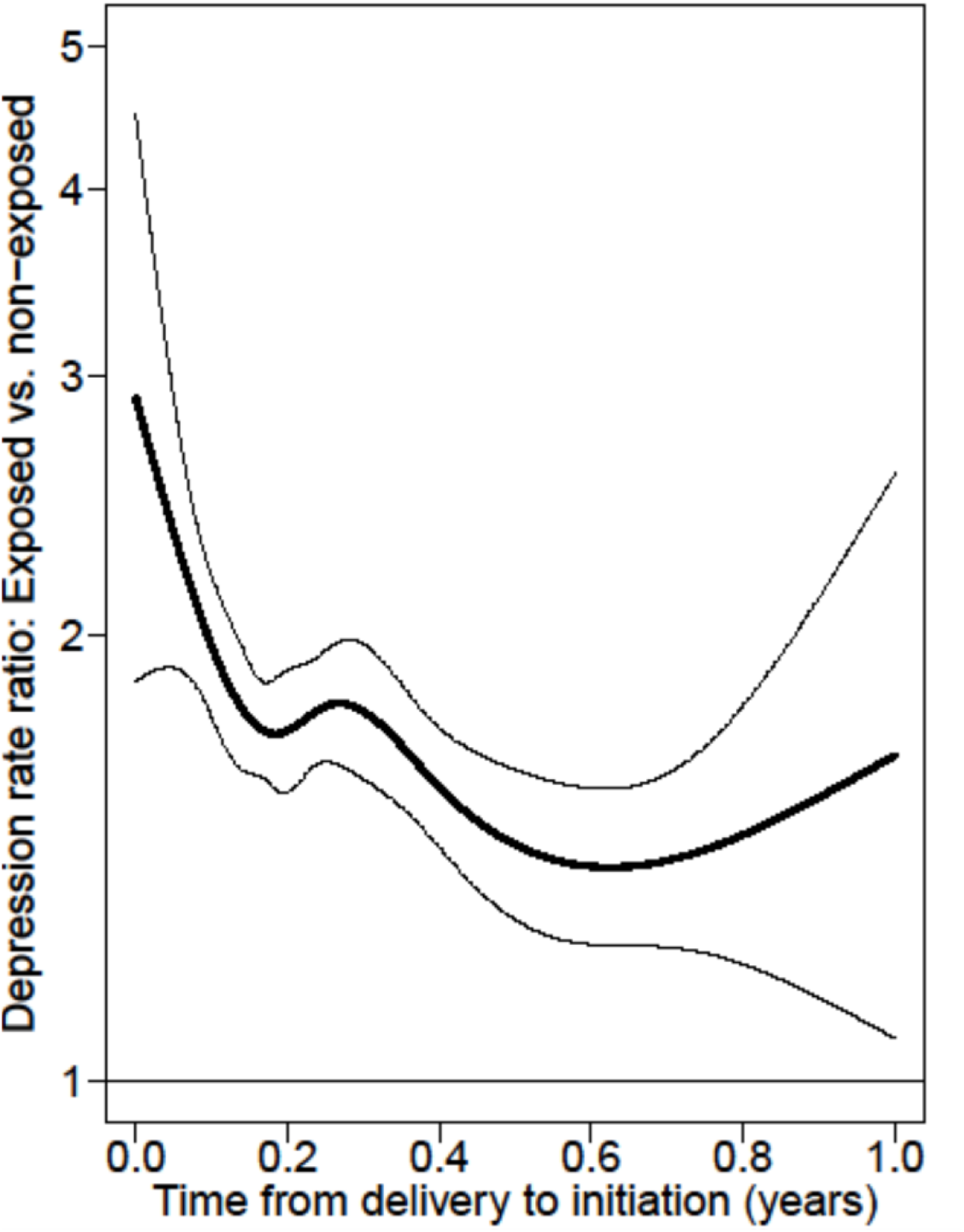
Estimated effect of time from delivery to initiation of combined oral contraception in exposed vs. non-exposed mothers in a model without the effect of time since initiation. The linear effect of time to initation in terms of rate ratios was estimated to 0.61 (95% CI, 0.48;0.79).

## Discussion

This study showed that HC initiation postpartum was associated with an increased risk of depression one year postpartum across all age groups. This phenomenon was most pronounced for women with no prior mental disorder. The risk was increased for COC, CNOC, and PNOC, but not for POP exposure.

Previous research on the association between postpartum HC use and depression has shown conflicting results.^7^ An observational study based on military healthcare system data found an increased risk of antidepressant use among women using subdermal implant and vaginal ring, no risk among COC or LNG-IUS users, and a reduced risk among POP users.^27^ As this study only followed POP users for 4.3 months on average, it may have missed essential follow-up time; we observed a time-varying POP effect across 12 months postpartum, which may reflect a delay between filling a prescription and actual time of initiation. Mothers in both studies filled a POP prescription shortly after delivery, however, other data have shown that despite having an early postpartum POP prescription, only about half of new mothers use them three and six months postpartum, possibly because they have not started them yet.^28^ Furthermore, the estimated risk can be attenuated by early discontinuation or even a lack of initiation among those who filled a prescription. In Denmark, as many as one out of four filled only one POP prescription.^3^ In addition, the lower risk in the early postpartum phase may be explained by selection bias; progestogen-only contraceptives are recommended over combined hormonal contraceptives while breastfeeding due to a putative negative impact on lactation,^15^ hence, mothers who filled a POP prescription early postpartum might be overrepresented by breastfeeding mothers. Accordingly, 13% of POP users switch to COC within 12 months postpartum, which may reflect a switch after termination of breastfeeding.^3^

Time-varying effects may also occur if the risk is moderated by timing of initiation as shown for COC where the sooner the initiation after delivery the higher the depression rate. The early postpartum period may be a window of maternal vulnerability where the interplay between the abrupt hormonal changes and psychological stressors may moderate the risk related to HC use. Alternatively, it may also be explained by selection bias as those who initiate COC early after delivery may represent a selected group of non-breastfeeding mothers or mothers with a medical indication for choosing COC over the recommended progestogen-only products.

Two studies with more precise track of time of initiation have supported a link between postpartum progestogen exposure and the development of depressive symptoms; a double-blind, randomized placebo-controlled study showed increased risk of depressive symptoms in women allocated to a single depot injection compared to placebo at six weeks, but not three months postpartum;^29^ and a randomized study found more depressive symptoms reported at one and three months postpartum in women allocated to depot injections compared to women allocated to cobber IUS.^30^

The higher depression risk associated with HC exposure in women without, compared to women with a prior mental disorder, is supported by previous observational studies.^24,25,31^ However, this is in contrast to a randomized study, where prior or ongoing mental disorders were a risk factor for HC-induced mood lability.^32^ This discrepancy may be explained by a healthy user bias in the observational studies, i.e., women with a prior mental disorder may be less likely to be prescribed HC due to reported mood-related side effects. Alternatively, it may reflect that the relative contribution from HC exposure in the development of clinical depression plays a less prominent role relative to other risk factors in a high-risk group. Importantly, it shows that HC-associated depression is seen despite no prior mental disorder, which is also supported by a prospective study demonstrating postpartum mood symptoms associated with HC exposure irrespective of psychiatric symptoms during pregnancy.^33^

In contrast to outside the postpartum period,^4–6^ the current study found no conclusive pattern of higher risk associated with HC use among the younger relative to older women after stratifying on covariates showing evidence of time-varying effects. It has been hypothesized that a younger brain under development, i.e., in adolescence, may be more susceptible to exogenous hormones,^24^ but such difference in susceptibility may attenuate due to the structural and functional brain changes happening in relation to pregnancy and childbirth.^34^

The higher risk of depression in the postpartum period associated with HC initiation highlights the importance of considering contribution from HC exposure to the already heightened risk of depression in women in the postpartum period. Furthermore, providing that our explorative analysis is replicated, the timing of initiation may also be important to consider since the early postpartum period may represents a relevant window of vulnerability.Especially, this should be considered at postpartum contraceptive counseling where a history of HC-associated mood deterioration, premenstrual dysphoric disorder, or postpartum depression may add to such risk profiling.^35,36^

### Strengths and limitations

The strength of the study is the use of national health registers, providing a nationwide, unselected study population with information on various potential confounders. Further, it involves a population of women who have all just given birth for the first time and who were advised to consider contraceptive methods at the postpartum counselling. This approach may reduce some of the potential confounders related to the decision to start HC, which might be more significant at other lifetime periods.

Our study also has several limitations. First, the study is observational, hence, a causal link cannot be definitely inferred. Second, depression is not always the indication for antidepressant use which may lead to a misclassification bias; however, as many as 80% of antidepressants are prescribed for depression during pregnancy,^37^ and if such a misclassification is expected to be non-differential, it would bias the results towards the null.^38^ Third, the day a prescription was filled may not mirror the day of initiation nor if HC was actually used postpartum, potentially leading to a misclassification which could bias towards the null. Fourth, the findings may also be attenuated by a healthy user bias due to women not starting HC postpartum because of previous adverse experiences. Last, our findings may not extend to milder cases of depression, which go undetected or are just not treated medically or diagnosed in any in or outpatient clinic.

## Conclusions

HC use was associated with an increased risk of depression in the postpartum period. This was observed for COC, CNOC, PNOC, but was inconclusive for POP. We observed no strong evidence of an age effect on the HC-associated depression risk. These findings raise the question if the incidence of depression postpartum may be inflated by routine HC initiation, which is important information to convey at postpartum contraceptive counseling.

## Data Availability

Danish national health register data cannot be
distributed, but access to the data can be granted by the appropriate authorities

## Author Contributions

*Concept and design:* All authors

*Acquisition, analysis, or interpretation of data:* All authors

*Drafting of the manuscript:* Larsen.

*Critical revision of the manuscript for important intellectual content:* All authors.

*Statistical analysis*: Larsen, Ozenne.

*Obtaining funding*: Frokjaer.

*Administrative, technical, or material support*: Mikkelsen, Lidegaard.

*Supervision*: Ozenne, Mikkelsen, Liu, Bang-Madsen Lidegaard, Frokjaer.

## Non-Author Contributions

### Data Access, Responsibility, and Analysis

SV had full access to all the data in the study and takes responsibility for the integrity of the data and the accuracy of the data analysis.

### Paper presentation information

Abstract was presented at the 36th ECNP Congress, The European College of Neuropsychopharmacology, Barcelona, Spain, 7-10 October 2023.

