## Supplementary material for "Postpartum hormonal contraceptive use and risk of depression"

### Supplementary Online Content

**eMethods 1.** The multistate Markov cox models

**eMethods 2.** Flexible parametric multistate model with multiple timescale

**eFigure 1.** Study population flowchart

**eFigure 2.** Multistate models

**eFigure 3**. Cumulative incidence curves for postpartum initiation of hormonal contraception

**eFigure 4.** Time-varying hazard and hazard ratio for POP exposure

**eFigure 5.** Hazard ratio of depression compared to non-users dependent on stratification of covariates showing evidence of violating the proportional hazard assumption

**eFigure 6.** Sensitivity analyses

**eFigure 7.** Risk curves for depression postpartum when proportional hazard assumptions were relaxed for the transition intensity from the exposed and the non-exposed state to depression state in the multistate Markov Cox model

**eFigure 8**. Depression rate in relation to time since delivery, time since initiation and time from delivery to initiation of combined oral contraception

**eFigure 9**. Depression rate at different timings of initiation of combined oral contraception postpartum

**eTable 1.** Overview of registers, variables, and codes

**eTable 2.** Instantaneous risk of depression postpartum stratified on age groups

**eTable 3.** 12-month average absolute risk of depression in the postpartum period when the proportional hazard assumption was relaxed

**eMethods 1.** The multistate Markov cox models

The average absolute risks were estimated by use of multistate models considering three (or more when the different HC types were considered) possible states (non-exposed, exposed, and depressive state) where transition hazards were estimated using Cox models and the average risk under current/observed HC use was obtained via the G-formula by evaluating the risk for each individual and averaging these risks (formula 12 in Cortese et al 2020).[1] The multistate Markov cox models included two Cox regression models; one modelling the transition intensity from the non-exposed to the exposed state, and one modelling the transition intensities to the depressive state from the exposed and non-exposed states. The key assumptions were (i) proportional hazard between the exposed and non-exposed states to the depressive state: $\alpha_{23}(t, X) = \alpha_{13}(t, X)e^{\gamma}$ (**eFigure 2A**) where X is the vector of covariates (ii) the hazard in the exposed state was not dependent on the entrance time to this state (Markov assumption). The follow-up time of the mothers who initiated HC was therefore split into two; time from delivery to start of HC exposure and time from HC initiation to depression or end of follow-up. Both models were adjusted for covariates listed in the main text, which were assumed to be time-invariant. The hypothetical risk had nobody initiated HC was obtained by setting the transition intensity from non-exposed to exposed to zero, which is only valid under the additional assumptions; 1) first-time mothers start HC for reasons unrelated to their risk of depression conditioned on the included covariates and 2) any prevention from HC initiation does not affect the direct transition from the non-exposed to the depressive state. When average risks were estimated for the different HC types, the multistate model was expanded to contain a state for each HC type for which a transition intensity was calculated (**eFigure 2B**). In the main analysis we model the HC effect under the proportional hazard assumption. To evaluate the risks without relying on the proportional assumption for the HC effect, a different baseline hazard was used for $\alpha_{13}(t, X)$ (non-exposed) and $\alpha_{23}(t, X)$ (exposed) instead of using regression coefficient for the exposure, so risks and risk curves could be estimated without relying on proportional hazards but while keeping common log hazard ratio coefficients for the covariates. The RiskIDM function was used for fitting the multistate Markov Cox modelling (https://github.com/bozenne/butils/blob/master/R/riskIDM.R).

1. Cortese G, Andersen PK. Competing Risks and Time-Dependent Covariates. *Biometrical Journal*. 2010;52(1). doi:10.1002/bimj.200900076

**eMethods 2.** Flexible parametric multistate model with multiple timescale

In the Markovian multistate model with the non-exposed, the exposed (a state for each hormonal contraceptive type), and the depressive states, only one time scale was used, i.e., time since delivery. However, the effect on depression risk may depend on the time since start of the intermediates state, i.e. time since initiation of a hormonal contraceptive type (a second time scale), but also on the time to the intermediate state. To address if the instantaneous risk of depression depends on whether you start early or late on hormonal contraception after delivery, we consider an alternative model able to consider multiple time scales (model 5 in Iacobelli and B. Carstensen 2013[2]).

We used Poisson regression where time was split into intervals of one month using the Lexis from the Epi package in R[3]. Smooth parametric functions were used to model the dependence on the different time scales with cubic splines with 6 knots (this was done for age, calendar year, time since delivery, time since initiation, and time to initiation). We only report the result for COC initiation since confidence intervals were very large and symptomatic of a too limited sample size for other HC types. Likelihood ratio tests (LRT) were used to assess a whether there is an effect of each time scale either as linear or non-linear (i.e., by comparing a model with no effect vs. linear effect vs. with splines for that timescale). Graphical representations similar to those of Iacobelli and B. Carstensen 2013[2] were used to display the fitted splines for each time scales separately and the modeled exposure effect depending of time since delivery and time since initiation. To facilitate the latter visualization, non-linear or linear effects were dropped from the model if the p-value for the corresponding LRT was above 0.05.


2. Iacobelli S, Carstensen B. Multiple time scales in multi-state models. *Stat Med*. 2013;32(30). doi:10.1002/sim.5976
3. Plummer M, Carstensen B. Lexis: An R class for epidemiological studies with Long-term follow-up. *J Stat Softw*. 2011;38(5). doi:10.18637/jss.v038.i05

**eFigure 1.** Study population flowchart

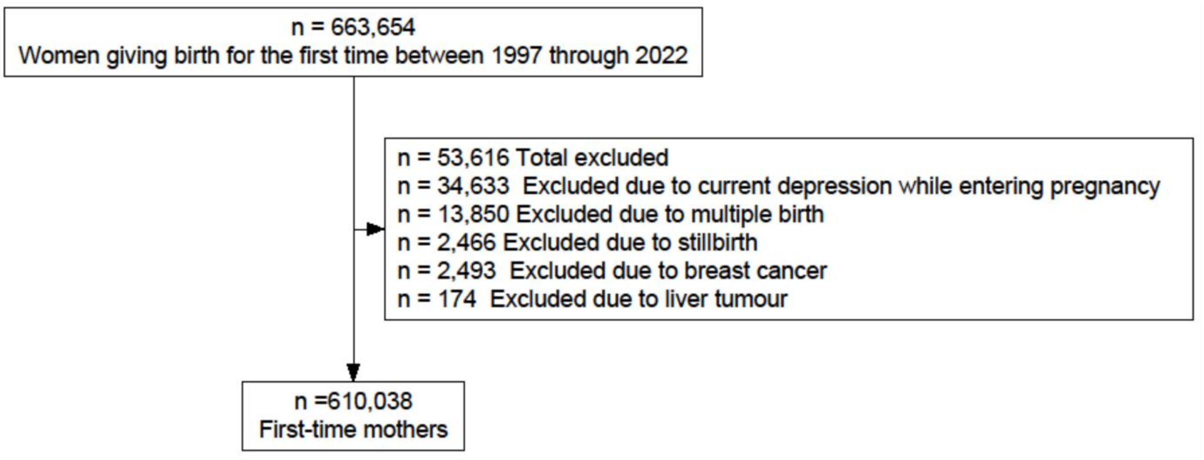


Flowchart illustrating the selection of the study population from the source population.

**eFigure 2.** Multistate models

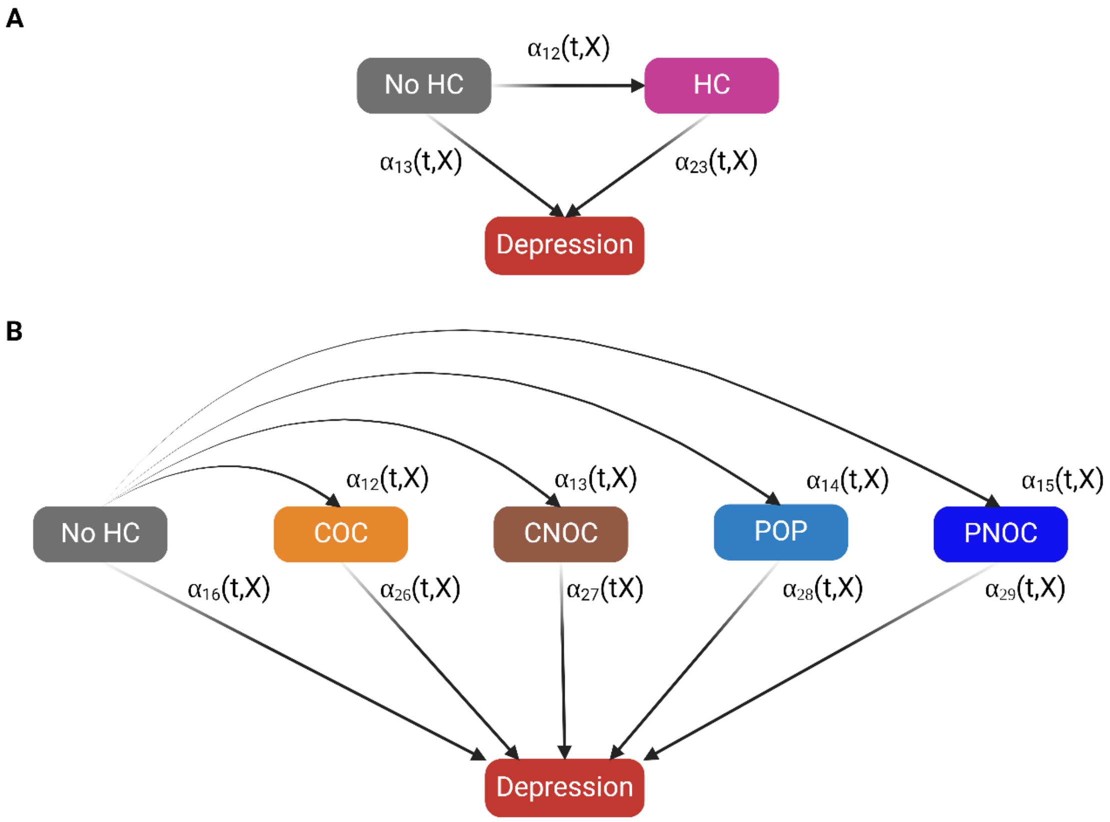


**A)** Multistate model with three possible states: "No HC", "HC", "Depression". $\alpha_{12}$, $\alpha_{13}$, and $\alpha_{23}$ refers to the transition intensities between the states. All women started in the "No HC" state and either stayed in this state or switched to the other states during the postpartum period. Mothers switched from non-exposed to HC exposed once they filled a HC prescription and stayed exposed during the rest of the follow-up time. **B**) This multistate model includes a state for each HC type grouped as "COC", "CNOC", "POP", and "PNOC". Once mothers initiated an HC type, they stayed exposed to this specific type for the rest of the follow-up time regardless of whether they discontinued or switched to another type.

HC, hormonal contraceptive. COC, combined oral contraceptive. CNOC, combined non-oral contraceptive. POP, progesterone-only pill. PNOC, progesterone-only non-oral contraceptive.

**eFigure 3**. Cumulative incidence curves for postpartum initiation of hormonal contraception

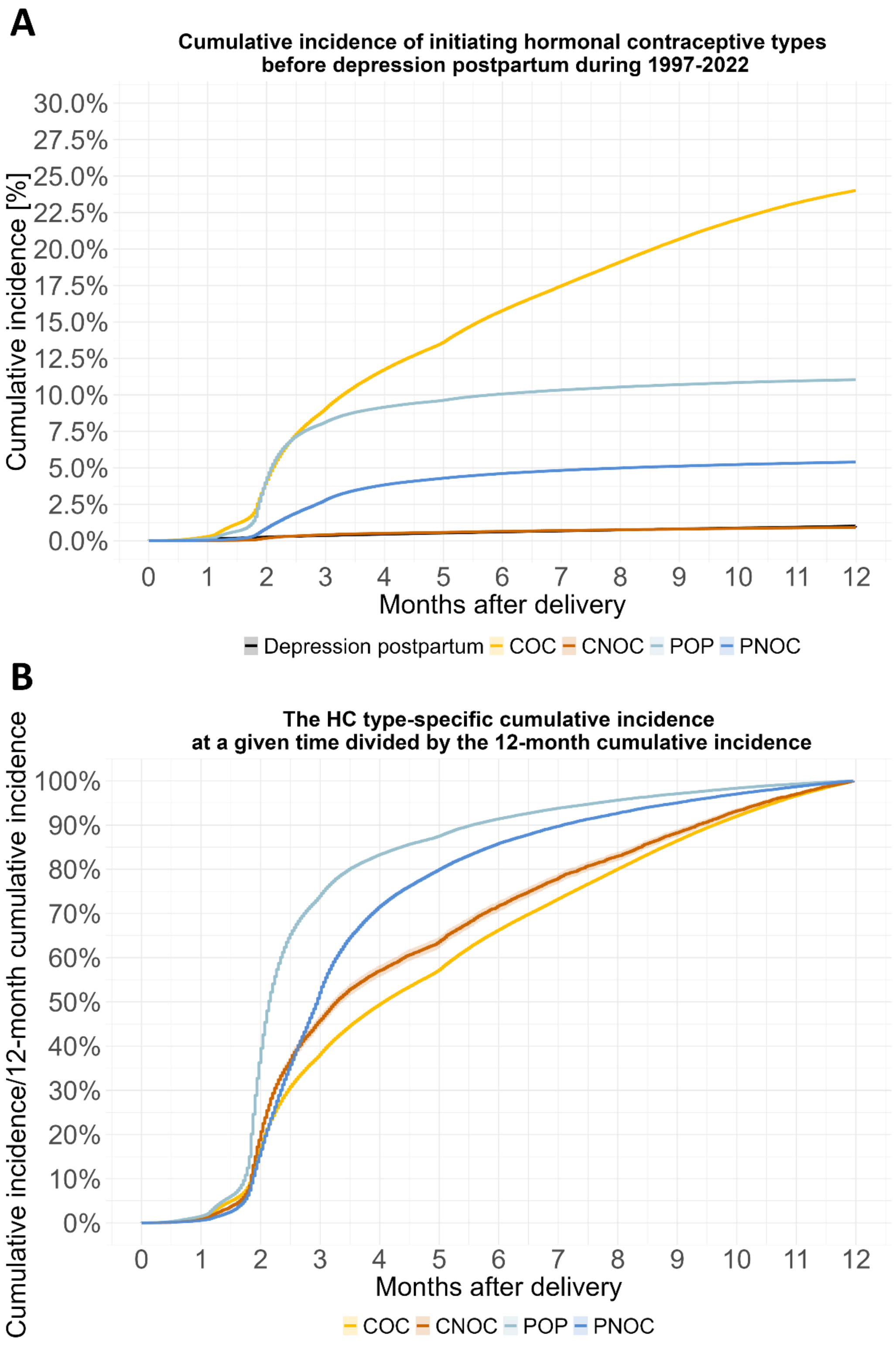


**A**) The cumulative incidence of initiating the different hormonal contraceptive types before depression postpartum. **B**) The cumulative incidence when divided by the 12 months cumulative incidence for each type.

The terminology "cumulative incidence" was used as the probability of starting contraception with depression postpartum as competing risk.

**eFigure 4.** Time-varying hazard and hazard ratio for POP exposure

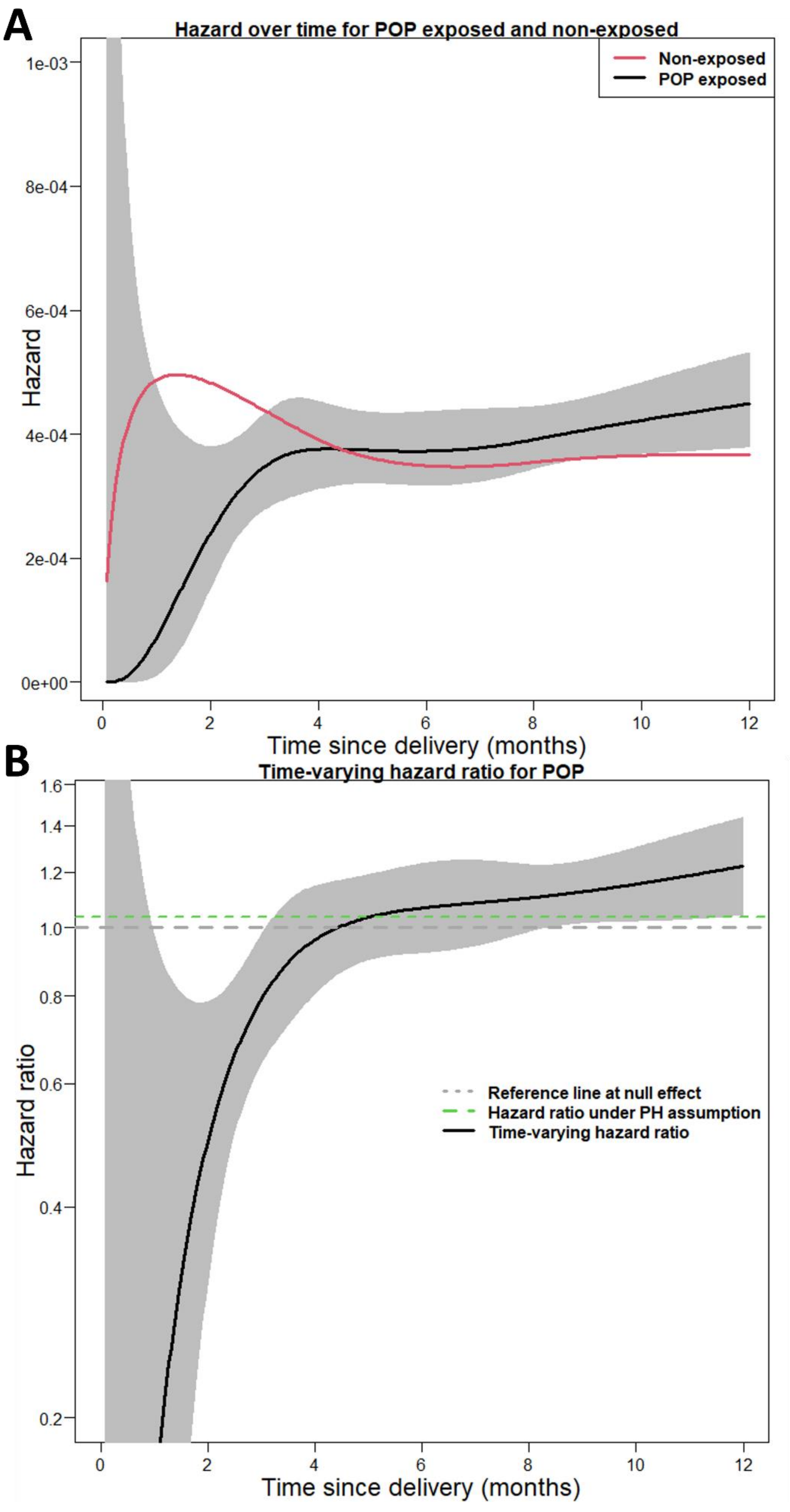

**A**) Time-varying hazard for developing depression in the postpartum period for mothers exposed to progestogen-only pill (POP) (solid black line with 95% confidence interval) and non-exposed (solid red line) across the postpartum period. **B**) The time-varying hazard ratio (HR) (the solid line with 95% confidence interval) for developing depression in the postpartum period for women starting on progesterone-only pills (POP) compared to non-exposed adjusted for age, calendar year, educational level, civil status, history of mental disorder, parental disposition for mental disorders, medical indications for HC use, IVF-treatment, preterm birth, instrument-assisted- or caesarean delivery, pre-eclampsia/eclampsia, pre-gestational- or gestational diabetes. Missing values were handled by imputing to separate groups for educational level, civil status, and preterm delivery status. The dotted gray line shows the HR=1 and the dotted green line shows the estimated HR under the proportional hazard (PH) assumption (HR=1.04 (95% CI, 0.95;1.13). The large reduction in the instantaneous risk in the early postpartum period is due to a relative high hazard for the non-exposed in combination with very few cases among the few mothers who had already filled a POP prescription shortly after delivery, hence the early postpartum HR has a very large uncertainty illustrated by the wide confidence interval.

**eFigure 5**. Hazard ratio of depression compared to non-users dependent on stratification of covariates showing evidence of violating the proportional hazard assumption

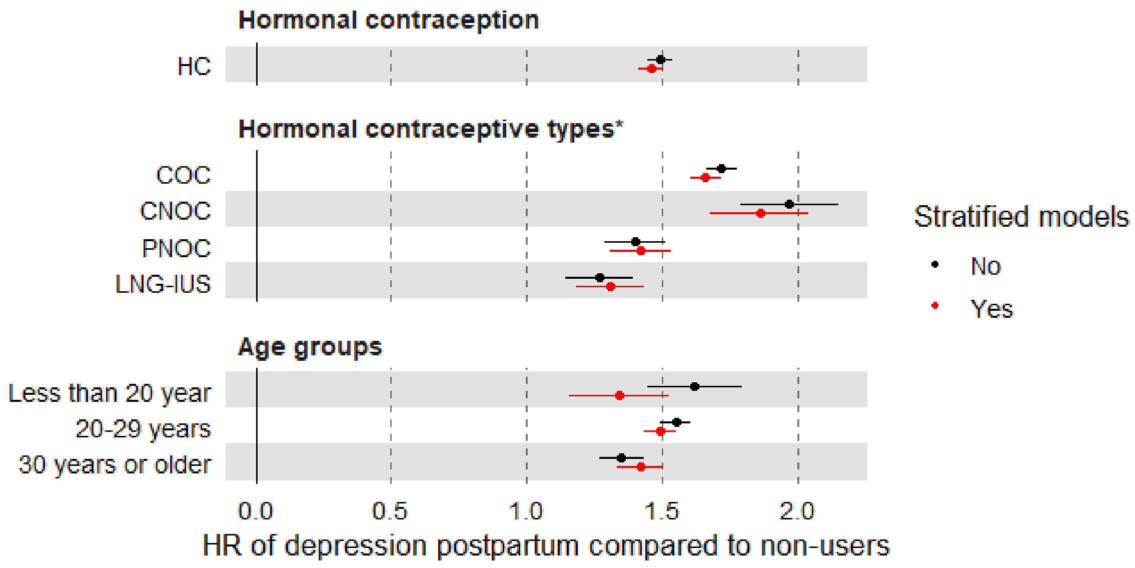


*HRs for progestogen-only pills violated the non-proportional hazard (Non-PH) assumption, hence the time-varying HR is shown in e**Figure 4**.

The Cox regression models were adjusted for age group, calendar period, educational level, civil status, history of mental disorder; parental disposition for mental disorders, medical indications for HC use, IVF-treatment, preterm birth, instrument-assisted- or caesarean delivery, pre-eclampsia/eclampsia, and pre-gestational- or gestational diabetes. The proportional hazard was relaxed for the following variables; age group, calendar period, educational level, civil status, and IVF-treatment. Missing values were handled by imputing to a separate group educational level (0.2% missing data) and preterm delivery status (0.1% missing data).

HC, hormonal contraceptive. COC, combined oral contraceptive. CNOC, combined non-oral contraceptive. PNOC, progesterone-only non-oral contraceptive. LNG-IUS, levonorgestrel-releasing intrauterine system

**eFigure 6.** Sensitivity analyses

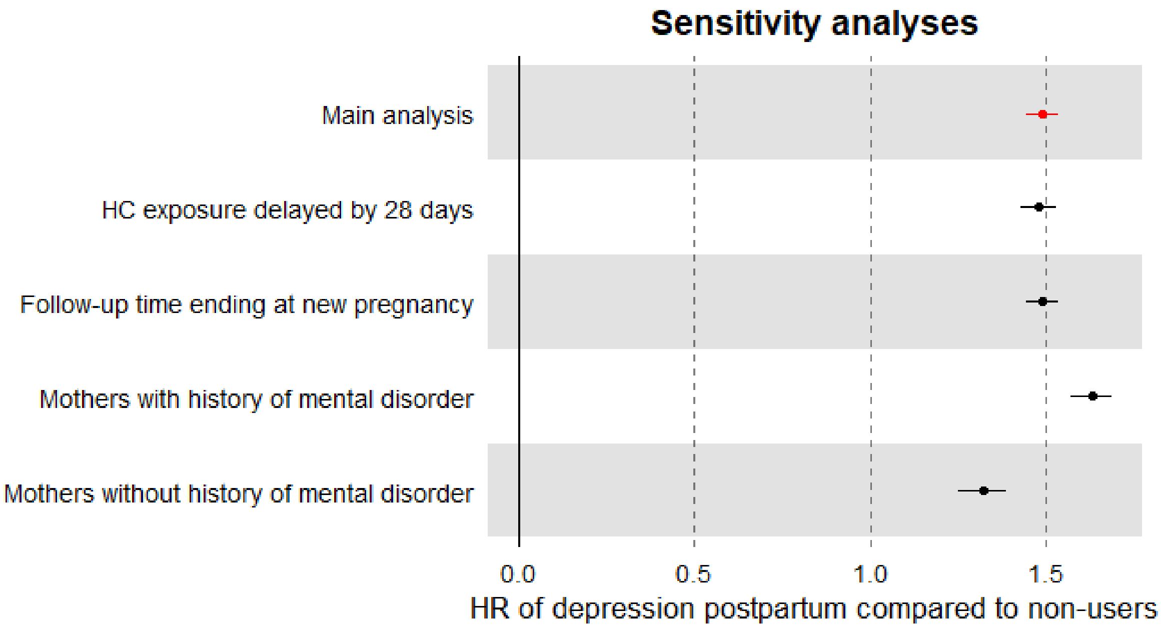


Hazard ratios are adjusted for age, calendar year, educational level, civil status, history of mental disorder*, parental disposition for mental disorders, medical indications for HC use, IVF-treatment, preterm birth, instrument-assisted- or caesarean delivery, pre-eclampsia/eclampsia, pre-gestational- or gestational diabetes. Missing values were handled by imputing to separate groups for educational level and preterm delivery status.

*The lower two analysis were stratified on prior mental disorder.

HC, hormonal contraceptive.

**eFigure 7.** Risk curves for depression postpartum when proportional hazard assumptions were relaxed for the transition intensity from the exposed and the non-exposed state to depression state in the multistate Markov Cox model

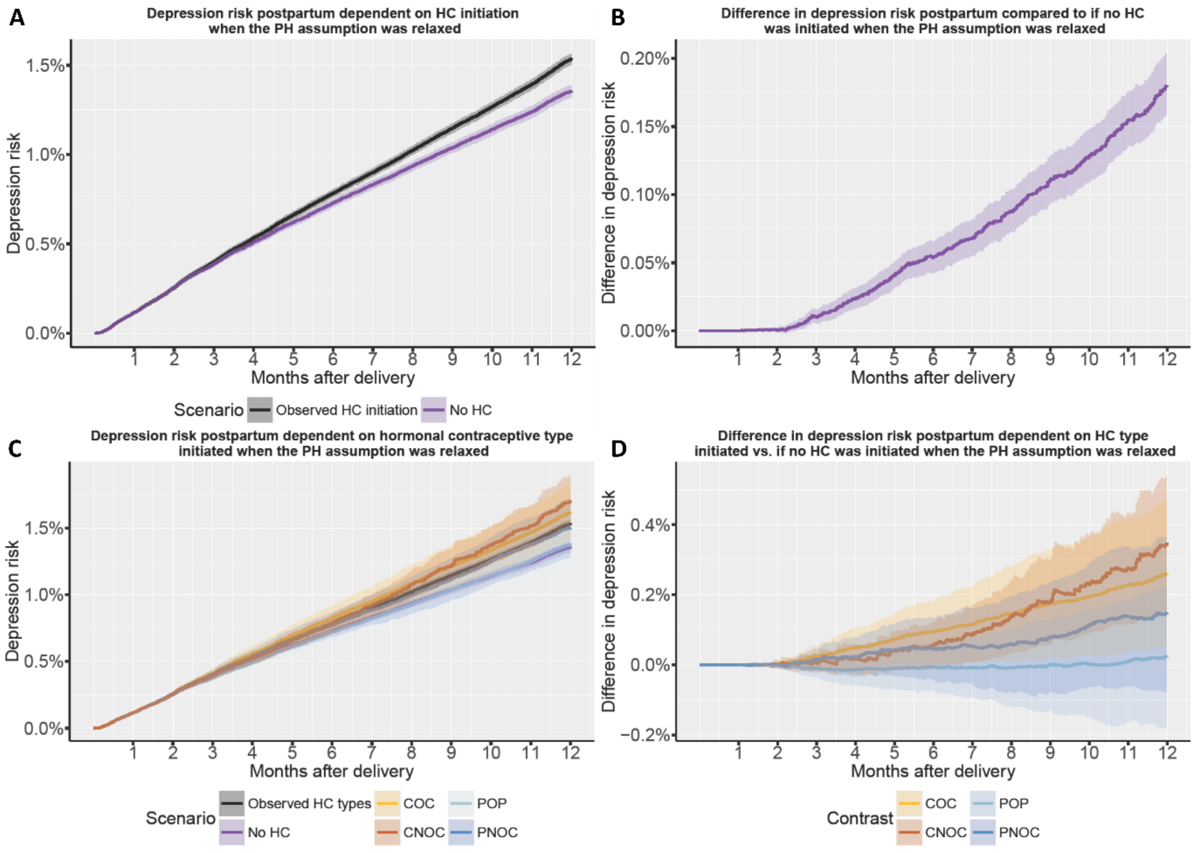


**A**) Shows the average absolute risk with 95% confidence intervals of depression within 12 months from delivery for the observed population with the observed fraction of mothers initiating hormonal contraception (HC) postpartum with the observed transition intensity vs. if nobody had initiated HC when the proportional hazard (PH) assumption was relaxed. **B**) Shows the average absolute risk difference with 95% confidence interval within 12 postpartum between these two scenarios when the PH assumption was relaxed. **C**) Shows the average absolute risk with 95% confidence intervals of depression within 12 months from delivery for the scenarios had all mothers who initiated HC initiated either combined oral contraception (COC), combined non-oral contraception (CNOC), progestogen-only pill (POP), or progestogen-only non-oral contraception (PNOC), with the observed transition intensity observed for each type vs. if nobody had initiated HC when the PH assumption was relaxed. **D**) Shows the average absolute risk difference with 95% confidence interval within 12 postpartum between these scenarios when the PH assumption was relaxed. The CNOC curve appears to increase more with time and crosses the curve for PNOC and COC, however, it is also estimated with a large degree of uncertainty due to the small number of CNOC users (n=5465, 0.9%), which is evident by the wide confidence intervals.

**eFigure 8.** Depression rate in relation to time since delivery, time since initiation and time from delivery to initiation of combined oral contraception

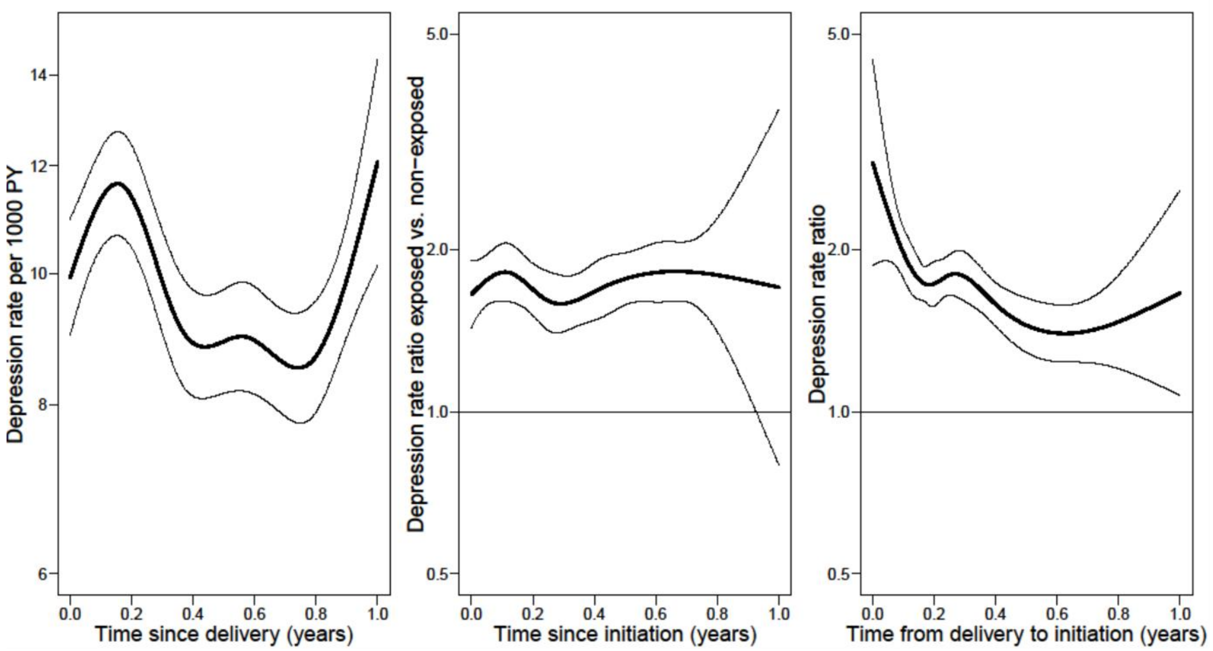


The graphs display estimated regression effects, i.e. effects varying with a specific timescale while all other covariates and timescale are being fixed, which is not realistic but provides insides into the model fit. The first panel shows the effect of time since delivery on the estimated depression rate per 1000 person-years for non-exposed mothers. The second panel shows the effect of time since initiation of combined oral contraception (COC) on depression rate between COC exposed and non-exposed mothers with same time since delivery and same time to initiation. Likelihood-ratio test showed no evidence of a linear effect vs. no effect (p=0.58) nor non-linear vs. linear effect (p=0.44) of time since initiation. The third panel shows the effect of time from delivery to COC initiation on the depression rate between COC exposed mothers with the same time since delivery and time since initiation (with reference to mothers starting 2 months after delivery). LRT showed evidence of a linear vs. no effect (p=0.000) of time from delivery to initiation of COC, but no evidence of an improvement of the fit from a linear to non-linear effect (p=0.13).

**eFigure 9**. Depression rate at different timings of initiation of combined oral contraception postpartum

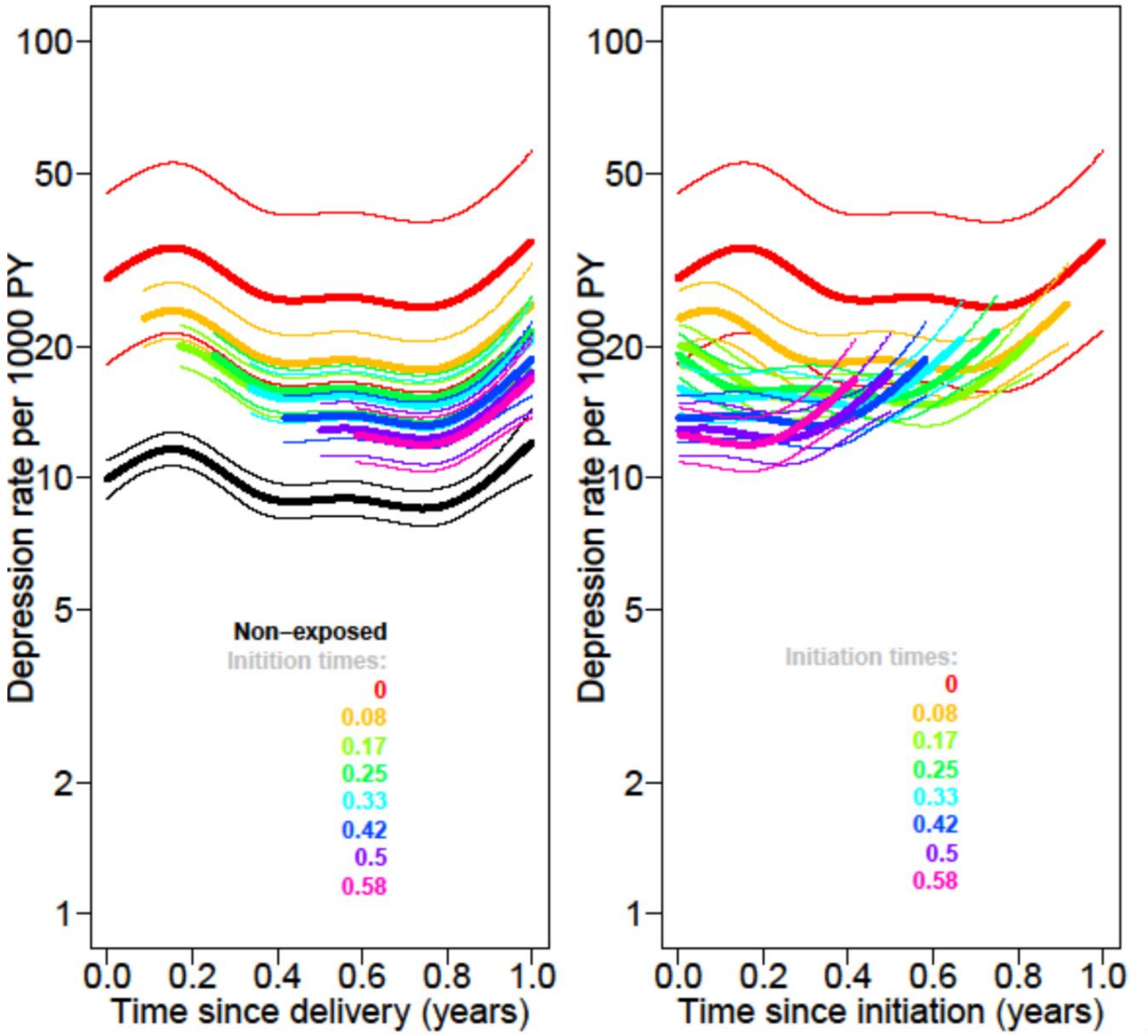


The estimated depression rate for non-exposed mothers and mothers initiating combined oral contraception at different time-points postpartum aligned on two time scales; time since delivery (left panel) and time since initiation (right panel). The rates are estimated while not including an effect of time since initiation of combined oral contraception. Estimates are represented with a thick line and 95% confidence intervals specific to each timepoint with thin lines. The rate ratio of the linear effect of time from delivery to initiation was 0.61 (0.48;0.79).

**eTable 1. Overview of registers, variables, and codes**

| Registers | Variables | Codes |
|---|---|---|
| **Danish Civil Registration System** (data since 1968) | | |
| | Date of birth, immigration, emigration, kinship, civil status | |
| **Demographic Registers of Statistics Denmark** | | |
| | Educational degree | |
| **Danish Medical Birth Register** (complete data since 1973) | | |
| | Twin birth, stillbirth, gestational age, smoking status | |
| **Danish National Patient Register** (complete data since 1977) | | |
| | Depression postpartum | ICD-8: 296.09, 296.29, 298.09, 300.49, 301.19<br>ICD-10: F32-34, F38, F39, F530 |
| | Instrument-assisted and caesarean delivery | ICD-10: O81, O82<br>NCSP-D: KMAE, KMAF0-2, KMAG03, KMAG13, KMCA |
| | Preeclampsia/eclampsia | ICD-8: 637, ICD-10: O11, O14-15 |
| | Pre-gestational/gestational diabetes | ICD-8: 249-250<br>ICD-10: E10-14, O24 |
| | Breast cancer | ICD-8: 174, 233<br>ICD-10: C50 |
| | Liver tumor | ICD.8: 155, 197.7-8,211.5, 230.5<br>ICD-10: C22, D015B, D134, D376A, C787 |
| Medical indications for HC use | Polycystic ovary syndrome | ICD-8: 256.9<br>ICD-10: E282 |
| | Endometriosis | ICD-8: 625.3<br>ICD-10: N80 |
| | Premenstrual syndrome | ICD-10: N943 |
| | Dysmenorrhea | ICD-8: 626.3<br>ICD-10: N944-946 |
| | Heavy menstrual bleeding | ICD-8: 626.2<br>ICD-10: N92 |
| | Hirsutism | ICD-10: L680 |
| | Acne | ICD-8: 706.1<br>ICD-10: L70 |
| **The Psychiatric Central Register** (complete data since 1969 on hospital admission and 1995 on outpatient contacts) | | |
| | Mental disorders | ICD-8: 300.0-315.0 (except 302.0 and 302.3)<br>ICD-10: F00-F99 |
| **Danish Prescription Register** (complete data since 1995) | | |
| Hormonal contraceptives | COC | ATC: G03AA* (except for G03AA13), G03AB*, and G03HB01 |
| | CNOC | ATC: G03AA13 and G02BB01 |
| | POP | ATC: G03AC* (except for G03AC06 and G03AC08) |
| | PNOC | ATC: G02BA03, G03AC06, and G03AC08 |
| | Antidepressant medication | ATC: N06A* |
| | Psychotropic medicine | ATC: N05* and N06* |
| | IVF-treatment | ATC: G03G*, G03DA04, H01CC01, H01CC02, L02AE01 |

ATC: Anatomical Therapeutic Chemical Classification system; ICD-8: International Classification of Disease and Health Related Problems, 8th revision; ICD-10: 10th revision, NCSP-D: Nordic Medico-Statistical Committee (NOMESCO) Classification of Surgical Procedures – Denmark.

| **eTable 2.** Instantaneous risk **of depression postpartum stratified on age groups** | | | | | |
|---|---|---|---|---|---|
| **Exposure** | **Person-years** | **No. of events** | **aHR[a] (95% CI)** | **aHR[b] (95% CI)** | **aHR[c] (95% CI)** |
| **<20 years** | | | | | |
| Non-exposed | 9025 | 263 | 1 (reference) | 1 (reference) | 1 (reference) |
| HC exposed | 5487 | 252 | 1.65 (1.38;1.96) | 1.62 (1.37;1.93) | 1.59 (1.34;1.89) |
| **20-29 years** | | | | | |
| Non-exposed | 247,364 | 3340 | 1 (reference) | 1 (reference) | 1 (reference) |
| HC exposed | 108,414 | 2257 | 1.61 (1.52;1.70) | 1.55 (1.46;1.64) | 1.51 (1.43;1.60) |
| **≥30 years** | | | | | |
| Non-exposed | 178,232 | 2377 | 1 (reference) | 1 (reference) | 1 (reference) |
| HC exposed | 43,027 | 762 | 1.36 (1.26;1.48) | 1.35 (1.24;1.47) | 1.33 (1.22;1.44) |

[a]Adjusted for age and calendar year.

[b]Adjusted for age, calendar year, educational level, civil status, history of mental disorder, parental disposition for mental disorders, medical indications for HC use, IVF-treatment, preterm birth, instrument-assisted- or caesarean delivery, pre-eclampsia/eclampsia, pre-gestational- or gestational diabetes. Missing values were handled by imputing to separate groups for educational level and preterm delivery status.

[c]Adjusted for age, calendar year, educational level, civil status, history of mental disorder, parental disposition for mental disorders, medical indications for HC use, IVF-treatment, preterm birth, instrument-assisted- or caesarean delivery, pre-eclampsia/eclampsia, pre-gestational- or gestational diabetes, immigration status, parental educational level, and smoking status. Missing values were handled by use of multiple imputations for educational level, parental education, smoking status, and preterm delivery status.

HC, hormonal contraceptive. aHR, adjusted hazard ratio.

| eTable 3. 12-month average absolute risk of depression in the postpartum period when the proportional hazard assumption was relaxed | | |
|---|---|---|
| **Scenario** | **Absolute risk[a] % (95% CI)** | **Absolute risk difference[a] % (95% CI)** |
| No HC initiation[b] | 1.36 (1.32;1.39) | Reference |
| Observed HC initiation | 1.54 (1.51;1.57) | 0.18 (0.16;0.20) |
| All initiating HC start on COC[c] | 1.62 (1.40;1.83) | 0.26 (0.05;0.47) |
| All initiating HC start on CNOC[c] | 1.70 (1.51;1.90) | 0.35 (0.15;0.54) |
| All initiating HC start on POP[c] | 1.38 (1.17;1.58) | 0.02 (-0.18;0.22) |
| All initiating HC start on PNOC[c] | 1.50 (1.28;1.72) | 0.14 (-0.08;0.37) |
| All initiating HC start on IUS[c] | 1.45 (1.22;1.68) | 0.10 (-0.13;0.33) |

[a]Adjusted for age, calendar year, educational, civil status, immigration status, history of mental disorder; parental disposition for mental disorders, medical indications for HC use, IVF-treatment, preterm birth, instrument-assisted- or caesarean delivery, pre-eclampsia/eclampsia, pre-gestational- or gestational diabetes. Missing values were handled by imputing to separate groups for educational level and preterm delivery status.

[b]A hypothetical scenario obtained by setting the transition hazard from non-exposed to HC exposed to 0.

[c]A hypothetical scenario obtained by setting the transition hazard from non-exposed to COC/CNOC/POP/PNOC/IUS by adding the transition hazard for each type and by setting the transition hazard to 0 for the rest of the types.

HC, hormonal contraceptive. COC, combined oral contraceptive. CNOC, combined non-oral contraceptive. POP, progesterone-only pill. PNOC, progesterone-only non-oral contraceptive. IUS, intrauterine system.
